# Interdisciplinary perspectives on multimorbidity in Africa: developing an expanded conceptual model

**DOI:** 10.1101/2023.09.19.23295816

**Authors:** Justin Dixon, Ben Morton, Misheck J. Nkhata, Alan Silman, Ibrahim G. Simiyu, Stephen A. Spencer, Myrna Van Pinxteren, Christopher Bunn, Claire Calderwood, Clare I.R. Chandler, Edith Chikumbu, Amelia C. Crampin, John R. Hurst, Modou Jobe, Andre Pascal Kengne, Naomi S. Levitt, Mosa Moshabela, Mayowa Owolabi, Nasheeta Peer, Nozgechi Phiri, Sally J. Singh, Tsaone Tamuhla, Mandikudza Tembo, Nicki Tiffin, Eve Worrall, Nateiya M. Yongolo, Gift T. Banda, Fanuel Bickton, Abbi-Monique Mamani Bilungula, Edna Bosire, Marlen Stacy Chawani, Beatrice Chinoko, Mphatso Chisala, Jonathan Chiwanda, Sarah Drew, Lindsay Farrant, Rashida A. Ferrand, Mtisunge Gondwe, Celia L. Gregson, Richard Harding, Dan Kajungu, Stephen Kasenda, Winceslaus Katagira, Duncan Kwaitana, Emily Mendenhall, Adwoa Bemah Boamah Mensah, Modai Mnenula, Lovemore Mupaza, Maud Mwakasungula, Wisdom Nakanga, Chiratidzo Ndhlovu, Kennedy Nkhoma, Owen Nkoka, Edwina Addo Opare-Lokko, Jacob Phulusa, Alison Price, Jamie Rylance, Charity Salima, Sangwani Salimu, Joachim Sturmberg, Elizabeth Vale, Felix Limbani

**Affiliations:** The Health Research Unit Zimbabwe, Biomedical Research and Training Institute, Harare, Zimbabwe; Department of Global Health and Development, London School of Hygiene and Tropical Medicine, London, UK; Malawi-Liverpool-Wellcome Programme, Chichiri 3, Blantyre, Malawi; Department of Clinical Sciences, Liverpool School of Tropical Medicine, Liverpool, UK; SHLS Nursing and Midwifery, Teesside University, Middlesborough, UK; Nuffield Department of Orthopaedics, Rheumatology and Musculoskeletal Sciences, Oxford University, Oxford, UK; Department of Medicine and Chronic Disease Initiative for Africa, Faculty of Health Sciences, University of Cape Town, Cape Town, South Africa; Malawi Epidemiology and Intervention Research Unit, Lilongwe, Malawi; College of Social Sciences, University of Glasgow, Glasgow, Scotland, UK; Department of Clinical Research, London School of Hygiene & Tropical Medicine, London, UK; Department of Population Health, London School of Hygiene & Tropical Medicine, London, UK; School of Health and Wellbeing, University of Glasgow, Glasgow, UK; UCL Respiratory, University College London, London, UK; MRC Unit The Gambia at LSHTM, Banjul, The Gambia; Non-communicable Diseases Research Unit, South African Medical Research Council, Cape Town and Durban, South Africa; School of Nursing and Public Health, University of KwaZulu-Natal, Durban, South Africa; Centre for Genomic and Precision Medicine, University of Ibadan, Ibadan, Nigeria; Department of Respiratory Sciences, University of Leicester, Leicester, UK; South African National Bioinformatics Institute, University of the Western Cape, Cape Town, South Africa; Department of Vector Biology, Liverpool School of Tropical Medicine, Liverpool, UK; Kilimanjaro Clinical Research Institute, Moshi, Tanzania; Department of Rehabilitation Sciences, The Kamuzu University of Health Sciences, Blantyre, Malawi; Department of Physical Medicine and Rehabilitation, University of Kinshasa, Kinshasa, Democratic Republic of Congo; Brain and Mind Institute, Aga Khan University, Nairobi, Kenya; SAMRC Developmental Pathways for Health Research Unit, University of the Witwatersrand, Johannesburg, South Africa; Health Economics and Policy Unit, The Kamuzu University of Health Sciences, Blantyre, Malawi; Department of Non-communicable Diseases, Ministry of Health, Lilongwe, Malawi; Musculoskeletal Research Unit, Translational Health Sciences, Bristol Medical School, University of Bristol, Bristol, UK; School of Public Health and Family Medicine, Faculty of Health Sciences, University of Cape Town, Cape Town, South Africa; Cicely Saunders Institute, Florence Nightingale Faculty of Nursing Midwifery and Palliative Care, King’s College London, London, UK; Makerere University Centre for Health and Population Research, Makerere University, Kampala, Uganda; Makerere Lung Institute, Makerere University, Kampala, Uganda; Department of Family Medicine, The Kamuzu University of Health Sciences, Blantyre, Malawi; Edmund A. Walsh School of Foreign Service, Georgetown University, Washington, DC, USA; Department of Nursing, College of Health Sciences, Kwame Nkrumah University of Science and Technology, Kumasi, Ghana; College of Medicine, University of Malawi, Blantyre, Malawi; Island Hospice and Healthcare, Harare, Zimbabwe; Malawi NCD Alliance, Malawi; Deanery of Clinical Sciences, College of Medicine and Veterinary Medicine, University of Edinburgh, Edinburgh, UK; Internal Medicine Unit, Faculty of Medicine and Health Sciences, University of Zimbabwe, Harare, Zimbabwe; Greater Accra Regional Hospital, Faculty of Family Medicine, Ghana College of Physicians and Surgeons, Accra, Ghana; Achikondi Women and Community Friendly Health Services, Lilongwe, Malawi; School of Medicine and Public Health, Faculty of Health and Medicine, University of Newcastle, Newcastle, Australia; Foundation President of the International Society of Systems and Complexity Sciences for Health; University of the Witwatersrand, Johannesburg, South Africa

**Keywords:** Multimorbidity, sub-Saharan Africa, global health, interdisciplinarity

## Abstract

Multimorbidity is an emerging challenge for healthcare systems globally. It is commonly defined as the co-occurrence of two or more chronic conditions in one person, but the suitability and utility of this concept beyond high-income settings is uncertain. This article presents the findings from an interdisciplinary research initiative that drew together 60 academic and applied partners working in 10 African countries to critically consider existing concepts and definitions of multimorbidity, to evaluate their utility and limitations, and to co-develop an context-sensitive, interdisciplinary conceptual framing. This iterative process was guided by the principles of grounded theory and involved focus- and whole-group discussions during a three-day concept-building workshop, thematic coding of workshop discussions, and further post-workshop iterative development and refinement. The three main thematic domains that emerged from workshop discussions were: the disease-centricity of current concepts and definitions; the need to foreground what matters to people living with multimorbidity (PLWMM), families, and other stakeholders; and the need for conceptual breadth and flexibility to accommodate the contributions of multiple disciplinary perspectives and heterogeneity within and between different African countries. These themes fed into the development of an expanded conceptual model that centres the catastrophic impacts multimorbidity often has for PLWMM, their families and support structures, for service providers, and for resource-constrained healthcare systems.

## Introduction

Multimorbidity – commonly defined as the presence of two or more chronic conditions in one person (1) – has been the focus of increasing attention over the last decade. While the bodies of literature were initially weighted towards high-income countries (HICs), multimorbidity has more recently been recognised as a global health challenge that may be especially detrimental in low- and middle-income countries (LMICs).(2–5) The health systems of many LMICs, including in Africa are donor-dependent, organised around single diseases, predominantly the infectious diseases including HIV and tuberculosis (TB) and, despite rapid rises in non-communicable disease (NCD) prevalence, have limited funding for these conditions.(3) This has led to a disparity in quality of care for people with NCDs and a failure to adequately integrate NCD care into programmes for people with chronic infectious diseases.(6)

An initiative facilitated by the UK Academy of Medical Sciences set high-level priorities for responding to multimorbidity in a global context.(2) and, in collaboration with the Academy of Sciences of South Africa, for sub-Saharan Africa specifically(7). Priorities include research into the patterning, burden, and determinants of disease ‘clusters’;(8–11) improving prevention and treatment of multimorbidity;(12,13) and the development of upstream health systems interventions and care models to better respond to the needs of people living with multimorbidity (PLWMM).(14,15) With current concepts and models of multimorbidity disproportionately based on biomedical research in the global North,(15) these initiatives emphasise the need for a more equitable, Southern-led response, facilitated by changes within the research ecosystem(2,7). This includes the development of South-South and North-South partnerships; cross-disciplinary and cross-sectoral collaborations;(7) and the inclusion of a greater range of perspectives – including those of the social sciences, PLWMM and affected communities, and frontline health workers – to co-develop more effective, equitable and context-sensitive interventions.(5,7)

Multimorbidity is currently challenging academics and practitioners working across Africa to step beyond entrenched disciplinary and disease ‘siloes’.(16) However, multimorbidity continues to mean different things to different stakeholder groups, reflecting the diversity of disciplines, perspectives, methods, measurement instruments, and geographic vantage points from which they enter this emerging field. Indeed, despite its apparent simplicity and widespread use, the definition of multimorbidity as ‘two or more chronic conditions’ endorsed by the WHO(1) and Academy of Medical Sciences(2) is not only contested but has also been heterogeneously interpreted in terms of which condition combinations ‘count’ as multimorbidity.(17,18) Consequently, it remains challenging to compare multimorbidity across datasets, to communicate across disciplines, and more broadly to ensure a coordinated response within and between African countries and regions. As commentators have noted, the multimorbidity conversation has become preoccupied with the promise of a universal definition, assumed to be a prerequisite for action, but has yielded poor returns.(19) Others have argued that the power of multimorbidity may lie precisely in its resistance to being pinned down to a number or specification of conditions, forcing us instead to consider the whole person in context.(16) This is a compelling proposition, one that speaks to the promise commonly pinned to multimorbidity for delivering on aspirations for holistic, person-centred care.(20) Yet without a common lexicon that enables communication across different disciplines, we may miss the opportunity to build on the current momentum building around multimorbidity to maximise benefits to patients and their carers across geographies, incomes and societal structures.

Responding to this need, this article presents the findings and outcomes from an interdisciplinary research initiative to interrogate the conceptual underpinnings of multimorbidity research and care in Africa. Grounded in a concept-building workshop in Blantyre, Malawi, the aim was to critically explore current definitions and concepts of multimorbidity; to critically appraise their potential, limitations, and utility; and to work towards a common conceptual model sensitive to the particularities and heterogeneity of African contexts. By folding a wide range of disciplinary perspectives, concerns, and interests into a common framework, the resultant multimorbidity model can, we contend, underpin and orient the cross-disciplinary, Southern-led response that, as described above, will be crucial to realize holistic, person-centred, and context-specific care for PLWMM.

## Methods

### Research Design

We used an inductive, co-productive research design. Guided by the principles of grounded theory,(21) the research process involved a three-day concept-building workshop in Blantyre, Malawi (June 22-24, 2022),(22) thematic ‘open coding’ of workshop discussions, and further iterative development of a conceptual model following the workshop. Our approach follows the growing interest in “collective experimentation”(23) to address issues of transdisciplinary concern in public and global health,(23–25) of which multimorbidity is arguably a paradigmatic example.

### Participants and Sampling

The workshop organising committee comprised an interdisciplinary group of public health researchers, clinicians, and social scientists (EB, CIRC, JD, RAF, FL, EM, BM). Potential participants were identified through purposive and snowballing methods between June 2021-March 2022, which are described in greater detail elsewhere.(22) The collaborator group drew together 60 researchers, clinicians, health planners, and policymakers (HIC-based n=19, LMIC-based (n=41), together representing a wide range of disciplinary perspectives, including from (sub-)fields of epidemiology, public health, clinical medicine, and the social sciences (Figure 1). Given the current concentration of multimorbidity research, and the location of the workshop in Malawi, the regional expertise among the collaborator group stemmed primarily from countries within Southern Africa, with the greatest concentration of experience from Malawi (n=33), South Africa (n=13), and Zimbabwe (n=9). Participants, several of whom were working across multiple countries, also brought experience from Central, Eastern, and Western Africa, together representing experience from 10 African countries (Figure 1). For practical and ethical reasons, we did not directly include PLWMM in the workshop; however, we asked collaborators, including social scientists and civil society and community representatives present, to represent learnings from their interactions to give an understanding of different patient perspectives. A detailed breakdown of participants’ institutional location, disciplinary and regional expertise, gender, and career stage, and a reflexivity statement detailing the measures taken to promote equitable partnership within this collaboration is provided in the <u>Supplementary Material</u>.(26)

**Figure 1.**
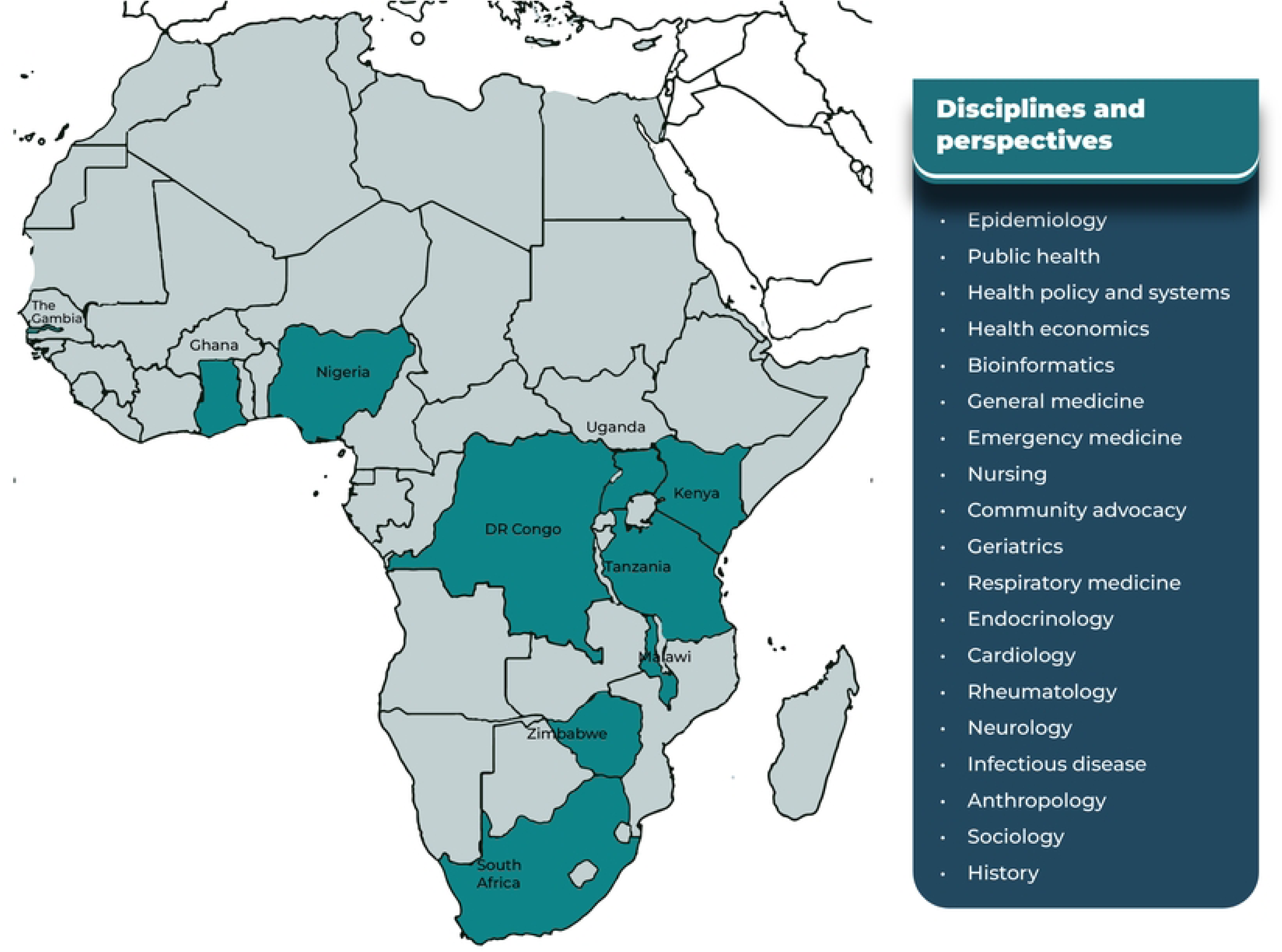
Regional and disciplinary expertise represented by workshop participants.

### Workshop design

A detailed account of the workshop design is published elsewhere.(22) The workshop was designed to optimise opportunities for cross-disciplinary discussion. Sessions were organised around four provisional thematic ‘domains’: (1) concepts and framings of multimorbidity; (2) population-level health data; (3) risk, prevention, and sites of intervention; (4) health systems and care models. Each session began with an ‘ignition’ talk which outlined current knowledge, gaps, and key questions within each domain. These questions were then addressed by participants through a combination of smaller focus groups aided by flip charts and plenary discussion. In a final session, reviewed and summarised the workshop sessions in plenary, collectively identifying core and cross-cutting themes, before breaking into working groups which advanced agreed themes for further development. This article was led by working group focused on developing a common concept of multimorbidity.

### Analytical framework

This research was guided by the principles of constructivist grounded theory, which holds that there is no ‘right or wrong’ and that emergent concepts and grounded theories are interpretive descriptions rather than an ‘objective’ account of reality.(21) Accordingly, the assumption underlying the workshop was that there is no privileged disciplinary vantage point from which to conceptualise or frame multimorbidity. Rather, different concepts and understandings foreground different aspects of the challenge multimorbidity presents for current systems, tied to particular knowledge bases, while potentially backgrounding others. This position, which was emphasised throughout the workshop and sustained through the post-workshop concept development, encouraged a diversity of concepts and understandings to be put into conversation, some of which diverged from and exposed the limits of the prevailing biomedical model. At the same time, in embracing difference, we were also able to find synergies and shared commitments. The embrace of both diversity and commonality formed the basis of developing a more cross-cutting, holistic, and ultimately more useful understanding of multimorbidity as it relates to African contexts.

### Data analysis

As this was an iterative, co-productive process, there was no rigid distinction between the process of ‘data collection’ and ‘analysis’.(21) All participants in the workshop engaged in critical analysis, many formed part of the core writing group, and all are recognised as authors. Source material for analysis included detailed notes of proceedings, including both focus groups and plenary discussions, taken by a team of rapporteurs (GTB, SS, IGS, SAS, NMY). Where necessary, the rapporteurs went back to individuals for clarification, which were absorbed into their notes; all notes were collated and reconciled to produce a unified account of workshop proceedings. Also included for analysis were the flip chart pages composed used during focus groups and the Microsoft Word documents produced using a shared screen during plenary discussion. A working group subsequently engaged in thematic open coding(27) of source documents on a shared drive to identify themes relating to the conceptualisation and framing of multimorbidity. Through regular analysis meetings, the working group iteratively worked towards themes of progressively higher orders of abstraction. The highest level of these themes, which we describe in the results section, were then fed into the development of an expanded model of multimorbidity. Refining this model was itself an iterative process, involving several iterations each of which was shared with the wider collaborator group along with the draft manuscript for further comment, discussion, and refinement.

### Research ethics

Due to the collaborative research design employed, in which all investigators were participants and vice versa, and all are named co-authors, formal ethical review was not required for this research. This follows recent examples adopting similar co-productive research models to develop cross-disciplinary frameworks and agendas.(24,28) All participants provided either verbal consent or written email consent to taking part in the concept-building workshop, which was captured using an Excel spreadsheet. At the end of the workshop, participants jointly agreed upon research outputs and working groups to take forward prominent themes from the discussions, and all consented to being named as co-authors on this particular output. All participants have reviewed the contents of the manuscript and have approved its final version. The concept-building workshop was subject to the safeguarding mechanisms of the host organisation (Malawi-Liverpool-Wellcome Programme), which included anonymous reporting procedures. We have not made available the detailed rapporteur notes as these would reveal the input of specific contributors. A detailed account of workshop proceedings, ratified and co-authored by all participants, has been published elsewhere.(22)

## Results

Three overarching themes that represent the group’s shared commitments for an African-context-sensitive conceptualisation of multimorbidity emerged: (1) the disease-centricity of prevailing concepts and definitions; (2) that models of multimorbidity need to be grounded in the realities and needs of PLWMM, families, and social networks; (3) the need for flexibility in defining multimorbidity for multimorbidity to be fit for purpose. These themes and their constituent sub-themes are summarised in Table 1.

**Table 1.** Key themes and sub-themes for conceptualising multimorbidity.

| Theme | Sub-Themes |
| --- | --- |
| Disease-centricity of current models of multimorbidity | • In medicine and global health, patients are defined by their diseases<br>• The minimalist definition of multimorbidity as two or more long-term conditions promotes a disease-centric view<br>• This definition also promotes narrow consideration of social factors associated with multimorbidity centred on individual behaviour and lifestyle factors<br>• Interventions working with this definition may continue to reinforce or exacerbate the status quo |
| Need to foreground what matters to PLWMM – their priorities, needs, and social context | • Benefit of focusing less on the disease categories within multimorbidity and more on its common consequences for PLWMM and associated needs<br>• The burden of multimorbidity on families, informal carers, and social networks is crucial to consider in many African contexts<br>• Need for a broader appreciation of the social, structural, and environmental context of multimorbidity<br>• Fragmented, disease-driven systems compound the burden of multimorbidity |
| Conceptual breadth and flexibility is needed for multimorbidity to be cross-cutting and useful | • A standard, one-size-fits-all definition not a panacea for multimorbidity<br>• Multimorbidity may mean different things, carry different priorities, and be more or less useful in different contexts and at different levels of scale<br>• A broad, flexible understanding that recognises multiple perspectives is needed to be cross-cutting and useful |

### Theme 1: Disease-centricity of current concepts

Across the countries represented at the workshop, a key challenge expressed by participants was that populations are living longer and facing increasingly complex disease burdens. Yet, health systems remain built around siloes of expertise based on single diseases. ‘Siloed’ approaches were observed to be perpetuated by ‘vertical’ funding models adopted by Northern donors, who have long prioritised acute diseases and chronic infectious diseases, notably HIV and TB (Box 1a). The single-disease approach, it was observed, drives many aspects of health systems from health policy and planning, to research and surveillance, to training and care delivery. It is central to the way health systems performance is monitored and evaluated and, in turn, why single disease care, especially care for those conditions prioritised by donors, continues to receive the majority of resources (Box 1b). Disease-centred thinking is so embedded that it is extremely challenging to step beyond this frame of reference; our systems seem to inevitably pull us back to a focus on diseases (Box 1c). As a result of the fragmentation of systems built around particular diseases, many areas of health including NCD and mental health are under-funded. Moreover, the needs of PLWMM are rarely considered within current systems. This is with the exception of integration of healthcare services for common comorbidities associated with HIV in some settings.

Throughout the workshop, participants grappled with the tension that, on the one hand, multimorbidity foregrounds disease concentrations and interactions rather than diseases in isolation. This makes it a potentially powerful concept for re-aligning priorities with the increasingly complex disease burdens affecting many African countries, including the colliding epidemic of communicable diseases and NCDs. On the other hand, the concept of multimorbidity, participants noted, is generally that of a compound disease category, most commonly defined as the presence or absence of two-or-more chronic conditions. As a result, it is still a disease-centred concept. Participants therefore questioned whether this definition was suited for moving us from a scenario in which people are defined by their diseases, to what was often referred to as a more ‘person-centred’ approach (Box 1d and 1e). Concerns raised about a disease-centred lens included that this may promote an overly simplistic, additive view of multimorbidity as the sum of interacting (but ultimately discrete) disease conditions. A related concern was the danger of emphasising the ‘communicability’ versus ‘non-communicability’ of diseases within multimorbidity. Continuing to reinforce this distinction within multimorbidity, it was noted, can perpetuate the tendency to reduce the social context of risk to ‘modifiable lifestyle factors’, a phenomenon that is particularly prominent in the context of NCDs (i.e., smoking, poor diet, sedentism, and substance abuse). As several participants noted, targeting individual behaviour and lifestyles does not amount to a ‘person-centred’ approach and may lead to patient shaming and stigmatisation (Box 1f), whilst deemphasising structural factors such as chronic poverty that constrain people’s life ‘choices’ and behaviours.

#### Box 1. Illustrative excerpts and quotes

a. “Most of the chronic diseases in sub-Saharan Africa countries such as HIV and TB are managed through vertical programs, which inhibits care for multimorbidity” (Session 1, plenary, rapporteur notes)
b. “The disease-centric outcomes that drive healthcare systems currently would need to be overhauled in order to respond to multimorbidity.” (Session 5, plenary, rapporteur notes)
c. “The challenge is that the system we work in does not accommodate a transformation in thinking; it keeps bringing us back to a focus on diseases” (Session 1, plenary, rapporteur notes)
d. “The definition of two or more conditions may have limitations, and may continue to perpetuate a disease specific approach” (Session 5, plenary, Word document on shared screen)
e. “We are still using a disease-focused lens to define multimorbidity – tensions versus a person-centred approach” (Session 5, group 5, flip chart excerpt)
f. “Labelling some of the non-communicable conditions related to multimorbidity, such as hypertension and diabetes, as ‘lifestyle diseases’ creates stigma and shaming” (Session 3, group 2, rapporteur notes)

### Theme 2: Need to foreground PLWMM’s priorities, needs, and social context

Continuous with theme 1, a particular concern expressed with the disease-based definition of multimorbidity is that it also fails to capture what *matters* to PLWMM. Several points about patients’ needs and priorities were highlighted. First, a concern voiced particularly by clinicians was that it often matters far less to people what the diagnoses and their causes are than their consequences and impacts, which become especially complex when it comes to secondary complications. Symptom and treatment burden, functionality, and quality of life were highlighted as important to fold into our understanding of multimorbidity (Box 2a), but it was noted that these are rarely considered within current disease-centred management approaches (Box 2b). While there may be considerable heterogeneity in the lived experience of multimorbidity across different condition combinations, it was also observed that the symptoms, needs and treatment burdens of PLWMM are often not disease-specific and may share similar profiles. Tellingly, when it was put to a vote whether the concept of multimorbidity should draw a distinction between ‘communicable’ and ‘non-communicable’ diseases, a significant majority said it should not. Participants were not suggesting that we cease thinking about causes and determinants of multimorbidity, which remain important especially within epidemiology, public health, and clinical medicine, but rather that we ground our concept of multimorbidity in the experiences and priorities of PLWMM.

Second, participants stressed that when considering the impacts of multimorbidity in many African contexts, it is vital to expand the focus beyond the affected person to the pivotal role that families, informal caregivers and larger support networks play in navigating burdens of illness and treatment and in PLWMM’s ability to ‘self’-manage (Box 2c). Accordantly, it was argued that while person-centred care is important, we need to be talking about *family*-centred care in this setting (Box 2d). A related consideration when designing multimorbidity care models for African health systems is not to ignore current social realities such as medical pluralism and the existing role of traditional healers. In the absence of such recognition, care models risk recreating Northern global health policies which fail to account for local experiences and African-centred knowledge systems and values.

Third, widening the lens further still, participants stressed that social, structural and environmental factors are crucial for understanding the patterning, impacts, and experience of multimorbidity. The concept of syndemics was recognised as a useful and well-established theoretical framework for capturing the bio-social causes and consequences of multimorbidity and how health and social challenges noxiously intersect (Box 2e). Social scientists reported from qualitative research that daily socio-economic struggles preceded and exacerbated bodily ailments and symptoms, and that within such fragile arrangements, interruptions to or changes in circumstance could have huge impacts on overall ability to cope, as was recently evidenced during the COVID-19 pandemic. Examples were given from different settings about the challenges PLWMM face: reaching secondary and tertiary health care when residing in rural areas; struggling to mobilise funds to spend on treatment and transport; and losing income when spending time at the clinic or balancing medical appointments with family responsibilities. It was further observed that specialised care organised around single diseases compounds the burden placed on those living with multimorbidity: it shapes which diseases are diagnosed and prioritised, the time and resources that are needed to (self-)manage different conditions, and the added care burden on families and carers (Box 2f and 2g).

#### Box 2. Illustrative excerpts and quotes

a. “The label of multimorbidity is likely to be useful if it includes burden and function, for instance pain, disability, and sleep” (Session 1, group 1, rapporteur notes)
b. “Clinics rarely account for patient preferences and needs. A patient-centred approach is where health care consciously works around patients’ needs, responding to individual preferences and trying to ensure that patient values guide clinical decisions.” (Session 4, ignition talk, rapporteur notes)
c. “Family members/community are very important for patients’ improvement because they have an influence on the treatment given, in the same way that nutrition post-delivery is mostly influenced by relatives” (Session 1, group 3, rapporteur notes)
d. “We need to think about family centred care – patient-centred care is important, but there is also a family who has to be involved in the process” (Session 5, plenary discussion, Word document on shared screen)
e. “Social determinants of health are important – syndemics, the commercial determinants of health, and others. For example, for TB patients, food insecurity, biological predisposition, and access to care have been used to identify patients/potential patients with multimorbidity” (Session 3, group 2, rapporteur notes)
f. “Most care in urban areas is specialized. This leads to fragmentation, adding a burden to the family and caretakers” (Session 1, group 3, rapporteur notes)
g. “Individual disease treatment compounds the burden. Patients start prioritising certain conditions and some may go untreated” (Session 1, group 2, rapporteur notes)

### Theme 3: Conceptual breadth and flexibility

The third theme was that, if context is taken seriously, a one-size-fits-all definition of multimorbidity trying to pin down multimorbidity to a specific number or combination of conditions is neither possible nor desirable (Box 3a). The diversity of academic and applied perspectives present at the workshop underscored how counterproductive, even harmful, a narrow definition favouring one discipline or perspective can be. The concept, it was agreed, needs to remain broad, flexible, and able to foreground different things depending on the question being asked, the problem or perspective driving the question, the geographical, epidemiological, and health system context, and indeed the level of scale. Depending on the context, multiple definitions may be needed (Box 3b).

Participants considered the utility of multimorbidity to different disciplinary and stakeholder groups, and how the priorities might shift in different contexts. As highlighted in Theme 2, the primary concern at the clinical and community level is functionality, quality of life, and the burden and complexity shouldered by PLWMM, their family and social networks (Box 3c). Public health researchers, while similarly recognising the need for greater emphasis on function and quality of life, were often more focused on the societal impact of multimorbidity, for which well-defined (though not necessarily disease-centric) measures are needed to facilitate comparison across datasets and to enable identification of patterns, trends, and burden (Box 3d). For policymakers and health planners, also deploying multimorbidity at the population level, the concept was viewed as useful for reconfiguring funding streams (e.g., from ‘vertical’ to ‘horizontal’ models), for developing new care delivery models (e.g., from disease- to person- and family-centred care), for training and deploying the health workforce, and managing risks among the population. Social scientists and historians pushed for an expansive concept, one connecting the intricacies of lived experience to the ‘upstream’ structural and systemic factors that socially pattern multimorbidity and exacerbate the burden (Box 3e). Finally, while PLWMM were not represented directly, the group noted that multimorbidity was unlikely to translate well into lay models (Box 3f), and that in fact this label could be harmful and stigmatising (Box 1f). Because of these different concerns, we may need to foreground (and background) different aspects of multimorbidity, and recognise scenarios when it is not useful, to maximise its potential and minimise its harms (Box 3g).

While recognising the impossibility of a narrow one-size-fits-all definition given that multimorbidity is, by nature, heterogenous and context-specific, the collaborator group was nonetheless optimistic about folding the multiple perspectives represented at the workshop into a broadly shared frame of reference to facilitate cross-disciplinary working. Following theme 2, it was proposed a focus on common consequences of multimorbidity and associated needs for PLWMM could be one pathway towards conceptual alignment. In the final session of the workshop, a working definition was proposed as: “A clustering of *needs* [added emphasis] and conditions that need to be addressed holistically, rather than in isolation”. Whilst only a starting point, it did draw together major points of agreement during the workshop: the foregrounding of clusters of needs rather than just medical conditions; the importance of a holistic purview; and the critique of compartmentalised approaches. Refining this concept was highlighted as a key aim moving forward.

#### Box 3. Illustrative excerpts and quotes

a. “There is no one-size-fits-all concept or framing of multimorbidity – context matters” (Session 5, group 1, flip chart)
b. “Some flexibility / ambiguity in definition – or multiple definitions – may be needed” (Session 5, plenary discussion, Word document on shared screen)
c. “For academics, policymakers, and public health, the label or definition of multimorbidity is likely to be useful, whereas, for patients, wellbeing and function are more important” (Session 1, group 1, rapporteur notes)
d. “Well-defined concepts are useful in academia. The label [of multimorbidity] is useful for prevalence/mapping clusters, being able to study interactions between drugs and conditions” (Session 1, group 1, rapporteur notes)
e. “You can’t do multimorbidity and focus only on medical conditions, there are a lot of things are going on – social, financial, medical. These factors work together and need to be addressed together, including clinical interventions, upstream solutions, downstream solutions, and community interventions.” (Session 4, ignition talk, rapporteur notes)
f. “It is important to distinguish the medical framework and the patient model. Biomedical and societal framings of symptoms translate poorly into lay terminology – there is not necessarily a term for multimorbidity” (Session 5, plenary discussion, Word document on shared screen)
g. “Part of the ‘art’ of multimorbidity may be centring diseases, people, and systems at different times and in different places and situations” (Session 5, group 1, rapporteur notes)

## Discussion

Thematic analysis of workshop proceedings revealed three core themes that are pertinent to the conceptualisation of multimorbidity in Africa: the disease-centricity of current definitions; the need to foreground what matters to PLWMM; and the importance of conceptual breadth and flexibility. Building on this framework, in the following, we place these core themes into conversation with current global health scholarship on multimorbidity, thereby developing an expanded model of multimorbidity that can underpin a coordinated but context-specific response to multimorbidity in Africa.

Our criticism of dominant constructions of multimorbidity emergent from Theme 1 resonates with several recent commentaries, reviews, and analyses. These have, in different ways, argued that a disease-centred framing of multimorbidity promotes a static, additive rendering of disease that treats illness as the sum of its parts.(16,20,29) One consequence of this disease-centred framing, as Blarikom et al.(16) have argued, is that the multimorbidity conversation has become preoccupied (and indeed, largely paralysed) by the search for standard biomedical definition and common core of conditions. Such a definition may be useful for drawing comparisons between populations and for identifying areas of met and unmet need. However, it may perpetuate a focus on diseases and individual behaviour and therefore ultimately fail to harness the critical potentials of multimorbidity to move beyond the single disease paradigm.(16,20,29) Against this backdrop, it is significant that attempts to adapt – and as one article puts it, “decolonise”(30) – multimorbidity for use beyond HICs highlight the need to move from a focus on diseases associated with ageing to recognise the ‘colliding epidemics’ of communicable and NCDs that characterise multimorbidity in LMICs.(3) While this may have important implications for integrating care across historically separated disease domains, the workshop proceedings suggest that ‘decolonising’ multimorbidity means more than adjusting its constituent diseases. Rather, beginning to decolonise the multimorbidity conversation means moving away from models of research and care that define and categorise people by their diseases.

The workshop concluded (as summarised in Theme 2) that there is a need to broaden our focus from the diseases that sit within multimorbidity to foreground what really matters to PLWMM in context. Calls for more holistic approaches resound in the multimorbidity literature from both HICs and LMICs and align with theoretical perspectives from multiple disciplines. This includes eco-social theory from epidemiology;(31) the syndemic framework,(32,33) the theory of recursive cascades(34) and burden of treatment theory(13) from the social sciences, and novel applications of complexity theory to multimorbidity within the primary care sciences.(29) Despite this wealth of theory and the recent proliferation of person-centred chronic care models,(14,35) in practice attempts to integrate care have not proven to be especially ‘person-centred’ in practice. Currently advocated-for person-centred care models are still generally designed using vertical disease programmes, for instance South Africa’s Integrated Chronic Disease Management (ICDM) system modelled on the country’s HIV programme.(15,36) Moreover, the common aim of such programmes to ‘activate’ PLWMM’s agency for better health decision-making emphasise individual responsibility, but often fail to respond to high rates of poverty and inequality that compromise the ability of PLWMM to control their exposure to health risks. A growing number of qualitative studies reporting PLWMM’s perspectives have been reported from countries including Malawi,(13) Ghana(37), Ethiopia,(38) and South Africa.(39) These show that PLWMM are unable to take control over their own health if they experience a ‘lack’ of health services, information, and basic necessities. Such lack in turn feeds into a cycle of precariousness that negatively impacts people’s ability to cope, often sending them down a slippery slope – or in complexity theory terms, over ‘tipping points’(29) – towards further disability and decline. Also appealed to but rarely prioritised in practice is the active involvement of caregivers and support networks in the provision of care for PLWMM.(15,40) Here, African social network theories such as Ubuntu – promoting mutual caring through compassion, reciprocity, dignity, and humanity – could be a useful concept, as they potentially cultivate resilience by creating a shared identity between PLWMM and carers, allowing them to flourish even when living in precarity.(40) Starting from where PLWMM are now – their priorities, values, needs, burden, and challenges navigating current healthcare systems – is a more promising way into the development of genuinely person-centred care models than the current retrospective ‘integration’ of different disease programmes. While an elaboration of what this might entail in practice is beyond the scope of this article, it would undoubtedly require careful reconsideration of what, where, how, and by whom care is delivered, likely involving the decentralisation of chronic care through healthcare workers trained in holistic, community-based care models – as elaborated, for instance, within the syndemic care framework.(33,41)

The main conclusion from Theme 3 was the need for a broad, flexible understanding of multimorbidity for it to be genuinely cross-cutting, folding in the possibility that in some scenarios it may not be so useful at all. This conclusion aligns with a growing body of multimorbidity scholarship running against the grain of the dominant drive to standardise and harmonise a biomedically-defined multimorbidity concept. A recent workshop exploring lessons from South Africa notably concluded that the ‘minimalist’ definition of multimorbidity as two or more chronic conditions, while appealing for its simplicity, is too simple. Any definition, it was argued, needs to take into consideration the causes and consequences of multimorbidity and will need to remain flexible enough to be responsive to different research questions, (inter-)disciplinary contexts, and scenarios.(42) Building on these observations, our findings suggest that a focus on common consequences for PLWMM holds particular promise for bringing different perspectives together around a common frame of reference. If, indeed, the needs and treatment burdens of PLWMM are often not disease specific, such a disease- agnostic focus could bring together disciplines and specialities in a way that has eluded the abstract and seemingly irresolvable attempts to pin down multimorbidity and its causal pathways in pathophysiological terms. The broader point, perhaps, is that the distinction between causes and consequences itself begins to break down once we recentre what matters to PLWMM, from whose perspective ‘consequences’ of a living with MM today may be ‘causes’ of further problems tomorrow.(34) By centring PLWMM and their ability to lead healthy, fulfilling and independent lives, we may better align medical and lay models of multimorbidity, such that it may not only be more cross-cutting and useful but also, perhaps, carry fewer negative connotations as a label. Figure 2 presents an expanded model of multimorbidity that folds in Themes 1, 2 and 3.

**Figure 2.**
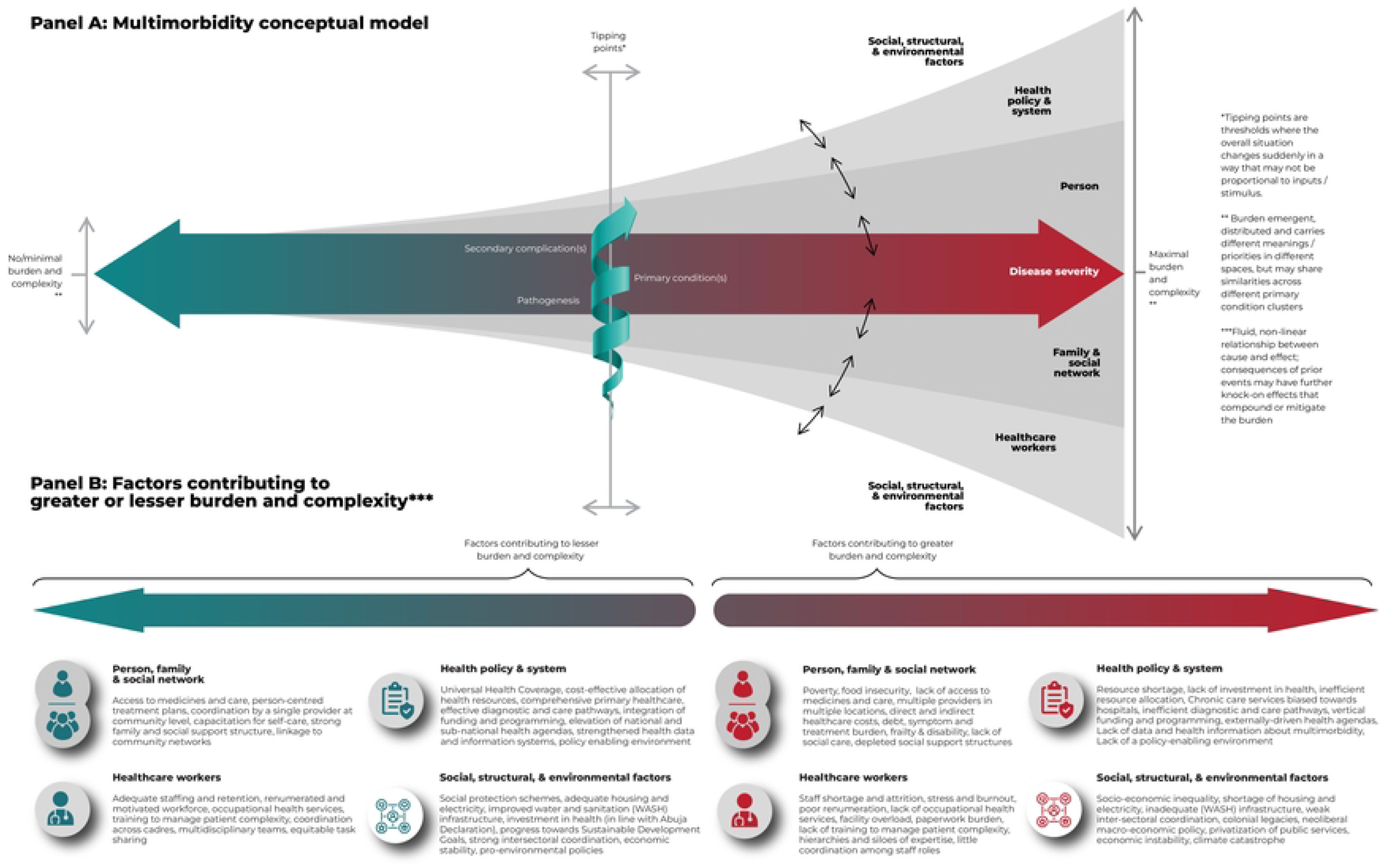
An expanded conceptual model of multimorbidity.

This model draws on the multiple theoretical influences of the participant group, including the syndemics framework,(32,33) burden of treatment theory,(13) and complexity theory.(29) It retains within it the basic idea of multimorbidity as involving more than one primary condition, in recognition of the utility of diagnostic categories within public health, epidemiology, and clinical medicine. But it remains agnostic about which kinds and combinations of condition ‘count’ as multimorbidity, instead expanding and bringing into the foreground the consequences of multimorbidity and its distributed burden on PLWMM, families, social networks, health providers, and the healthcare system. The model highlights factors identified by the collaborator group that currently overwhelmingly pull towards the right of the model, that is, that set PLWMM on a slippery (but not inevitable or linear) slope towards secondary complications and potentially catastrophic burdens that are felt across the system. It also casts our attention to the factors that might pull back people towards the left of the model. This includes seemingly small changes to a person’s circumstances that may make multimorbidity more manageable for the person affected and their wider networks, even in cases of advanced or complex disease. Given that many secondary complications cannot be reversed, the model suggests the emphasis should be on primary and secondary prevention. More than pharmacological or lifestyle interventions, this demands and rather interventions that take into account the interdependency between PLWMM and family, social network, healthcare providers, the health system, and wider social, structural, and environmental context.

While the ‘maximalist’ orientation of this multimorbidity model may lack some of the appeals of a minimalist definition, its dynamic and multidimensional nature enables different questions and disciplinary perspectives to be brought to bear on multimorbidity, including more refined definitions suited for specific questions. At the same time, the model suggests an overarching shift in the kinds of question that we ask and how we answer them. First, a focus on the complex bio-social interactions and impacts of multimorbidity moves us away from the disease-centred, cross-sectional designs from which most knowledge about multimorbidity to date originates (and that have tended to favour pharmacological and behavioural interventions) to disease-agnostic, richly contextual cohort research designs focused on burden, outcomes, and quality of life across the life course. Second, recognition of the multilevel factors at play implies that multimorbidity cannot be fully appreciated through any one disciplinary lens alone. Inter- and trans-disciplinary approaches, as well as holistic stakeholder engagement are needed to understand how biological, socio-cultural, political, economic, and environmental factors intertwine to co-produce multimorbidity; as well as to design holistic, person-centred, and systems-directed interventions. Third, the model implies a more context-specific approach to decide which disciplines and fields of expertise are relevant within the multimorbidity arena. Multimorbidity may result in similar needs and treatment burdens across different condition clusters, but its manifestation may also vary considerably across African settings given the tremendous heterogeneity of social, structural, environmental, and health system contexts. Thus, the knowledge bases and skillsets required to understand and address multimorbidity need to be similarly adaptive.(36) Whilst challenging, this is precisely the kind of context-sensitive approach that multimorbidity compels, offering a more promising pathway towards holistic person-centred care than universal, disease-centred interventions.

This model and the approach underlying it have several strengths. This was the first workshop to systematically bring together a multidisciplinary group of academic and applied actors to critically consider the meaning and utility of multimorbidity specifically within African settings. This was a bold exercise, not only in multidisciplinary experimentation,(23) but in elevating perspectives from a range of African contexts to reframe the multimorbidity conversation. This work provides a conceptual infrastructure to undergird the North-South and, more importantly, South-South partnerships that have been explicitly recognised as necessary for responding to multimorbidity in the region.(7) This research also had several limitations. First, while the collaborator group advocated for a person-centred perspective on multimorbidity, we were unable to directly include the voices of PLWMM. Further research is needed to gauge the extent to which this model resonates with PLWMM, whose perspectives remain under-represented within the multimorbidity conversation in Africa. The collaborator group was also biased towards Southern Africa, with the largest representation of expertise from Malawi, Zimbabwe, and South Africa, and thus our work disproportionately reflects views from these countries. Finally, our thematic analysis may be biased towards the views of the core working group. Whilst we endeavoured to represent the wider group’s perspectives objectively through a rigorous thematic analysis of workshop proceedings, the fact that we were both participants (in the workshop) and observers (analysing proceedings) means that the model may be biased towards our own values and viewpoints.

## Conclusion

In this article, we have analysed focus-and whole-group discussions from a workshop on multimorbidity in African contexts, placed emergent themes into conversation with current thinking on multimorbidity, and developed an expanded model based on the groups’ common commitments. While recognising the crucial research that has preceded our work, we believe in the necessity of providing nuance to the available framings of multimorbidity, stressing the importance of understanding the lived experiences of people and their networks, and adding in the socio-economic complexities that impact PLWMM, providers, and the development of systems. Further conceptual and empirical work is needed to draw out the implications of this conceptual model for the heterogenous and multifaceted health systems in Africa. Also open for the test of empirical scrutiny is whether it will prove useful for orienting and mapping different strands of multimorbidity work across disciplines, projects, and interventions, and for contributing to an overall more joined-up response moving forward.

## Data Availability

A detailed account of workshop proceedings, ratified by all participants, has been published in Wellcome Open [https://wellcomeopenresearch.org/articles/8-110] The original rapporteur notes and other sources materials have not been made available as these would reveal the input of specific contributors, all of whom are named co-authors of the article.

## Acknowledgements

We are greatly indebted to members of the Africa Multimorbidity Alliance who took part in the concept-building workshop on which this article is based. We are grateful to the administrators and technicians at the Malawi-Liverpool-Wellcome Programme for their efforts in preparing and running the workshop on which this article is based, with particular thanks to Kate Mangulama. We finally wish to express gratitude to the many funders and institutions who supported the travel and attendance of participants.

